# Capturing hand use of individuals with spinal cord injury at home using egocentric video: A feasibility study

**DOI:** 10.1101/2020.08.24.20180828

**Authors:** Jirapat Likitlersuang, Ryan J. Visée, Sukhvinder Kalsi-Ryan, José Zariffa

**Author notes:** **<u>The name, mailing address, email address, and telephone number of the person to whom proofs and reprint requests should be addressed (corresponding author):</u>** José Zariffa, KITE, Toronto Rehabilitation Institute, University Health Network, Toronto, Ontario, M5G 2A2, Canada.

## Abstract

**Background:** Measuring arm and hand function in the community is a critical unmet need of rehabilitation after cervical spinal cord injury (SCI). This information could provide clinicians and researchers with insight into an individual’s independence and reliance on care. Current techniques for monitoring upper limb function at home, including self-report and accelerometry, lack the necessary resolution to capture the performance of the hand in activities of daily living (ADLs). On the other hand, a wearable (egocentric) camera provides detailed video information about the hand and its interactions with the environment. Egocentric recordings at home have the potential to provide unbiased information captured directly in the user’s own living environment.

**Purpose:** To explore the feasibility of capturing egocentric video recordings in the home of individuals with SCI for hand function evaluation.

**Study Design:** Feasibility study

**Methods:** Three participants with SCI recorded ADLs at home without the presence of a researcher. Information regarding recording characteristics and compliance was obtained as well as structured and semi-structured interviews involving privacy, usefulness and usability. A video processing algorithm capable of detecting interactions between the hand and objects was applied to the home recordings.

**Results:** 98.58±1.05 % of the obtained footage was usable and included 4 to 8 unique activities over a span of 3 to 7 days. The interaction detection algorithm yielded an F1-score of 0.75±0.15.

**Conclusion:** Capturing ADLs using an egocentric camera in the home environment after SCI is feasible. Considerations regarding privacy, ease of use of the devices and scheduling of recordings are provided.

## Introduction

The return of arm and hand function is of the highest priority for individuals with cervical spinal cord injury (SCI) [1]. An individual’s ability to perform activities of daily living (ADLs) with their upper extremity (UE) is a crucial indication of independence and reliance on care. Outcome measures that can accurately quantify hand function in a natural context are needed for assessing new rehabilitation interventions and examining the transition back into the community after injury.

Current technique for monitoring hand function at home are limited to self-report and questionnaires, which are known to be sensitive to biases [2-8]. Such restrictions limit our understanding of how changes in UE function, extracted through direct observation by a clinician or investigator in a controlled environment, are impacting activity and participation at home and in the community. In particular, the *performance* domain of UE impairment (i.e., the actual use of the limb, as opposed to the capacity to use it [9]) cannot currently be measured directly.

In order to solve this gap in research, multiple studies have proposed wearable sensor systems based on accelerometers or inertial measurement units (IMUs). The devices are ideal for the home setting as they are wearable, relatively small, and easy to wear. However, they primarily capture arm movements, such as in studies in hemiparetic stroke survivors for comparing movement in impaired vs. unimpaired arms [10, 11], and lack the necessary resolution to capture information about the hand. Although previous studies demonstrated a relationship between accelerometry measures and independence, a disconnect remains between measured capacity and performance [12]. Similarly in SCI, accelerometers have been used to measure wheeling movements, independence, and the laterality of the injuries [13-14] but still without directly revealing information about hand function.

To address this limitation, we have previously proposed a wearable system based on a first-person (egocentric) camera, where the user’s point of view is recorded [15-16]. Egocentric video is not only able to capture hand movement information but also hand posture and its interactions with objects or the environment.

Furthermore, computer vision techniques for extracting information about the hand from egocentric video are a rapidly growing area of research, though not in the context of rehabilitation. Technical challenges including hand detection (locating the hand in the image) as well as segmentation (separating the outline of the hand from the background of the image) are described in [17-27]. Egocentric vision research to further extract information relating to ADLs has also been attempted, specifically in activity recognitions and object detections in ADLs [21-24]. However, generalizability can be a challenge in such systems, given the large variety of activities and objects found in the community.

In our previous work [15-16], we investigated the problem of detecting interactions between the hand and objects in the environment using automated analysis of egocentric videos. Detecting use of the UE in ADLs (performance) was conjectured to have applications as an indicator of independence. “Hand-objection interaction” is defined here as when the hand manipulates an object for a functional purpose; for example, resting a hand on the object would not constitute an interaction. Thus, the system is a binary classifier (interaction vs. no interaction) applied to each video frame.

In [15] we proposed a system to detect hand-object interactions in egocentric videos and evaluated it with able-bodied participants. In [16], we expanded our hand-object interaction system to participants with SCI (our target clinical population). Nevertheless, a limitation of these previous studies is that they were based on recordings in a controlled environment, namely a home simulation laboratory.

The present work expands the scope of this work to egocentric recordings at home. A feasibility study focusing on recording in participants’ homes was conducted. In this study, we explored the possibility of using our egocentric system in the participant’s own home to analyze unscripted activities performed without an investigator present. We report on the successes and challenges of deploying an egocentric wearable camera at home, as well as the technical performance of the system.

## Methods

### Dataset and Participants

Three participants (n=3) with SCI participated in this study. The inclusion criteria for this study consisted of participants with SCI self-reporting an impairment in hand function. There were no restrictions placed on the level or severity of injury, the amount of time since injury or on the etiology of the injury. Before the recording, participants met with the investigators in person, along with their primary caregiver (if applicable). During this visit, two assessments indicative of upper limb function were administered by the investigators: the Spinal Cord Independence Measure III (SCIM) [4-5] and the Graded Redefined Assessment of Strength, Sensibility and Prehension (GRASSP) [28]. Table 1 summarizes the participants’ demographics and upper limb assessments. The level of injury and AIS grade were self-reported by the participants, while the GRASSP and SCIM were assessed.

**Table 1:**
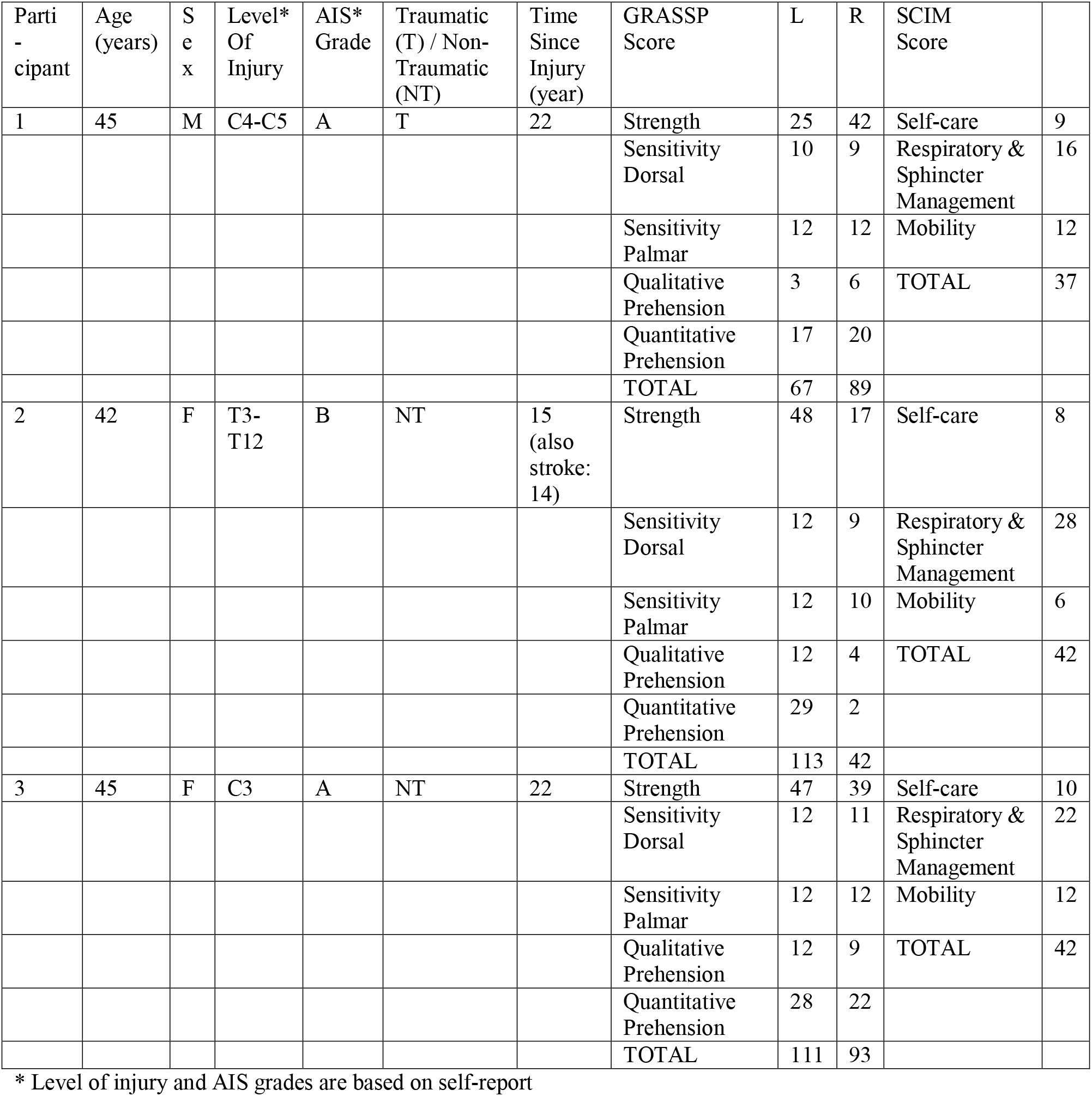
Participant demographics, injury characteristics, and assessment scores

Following a discussion with the participant, a schedule was made for video recordings in the participant’s home, without the investigators being present. They were asked to take part in three or more sessions of data recording of approximately 1.5 hours each over a two to three-week period. Egocentric video was recorded using a commercially available egocentric camera (GoPro Hero5, San Mateo, California, USA) worn by the participant on a head strap. A caregiver could help to set up the camera, if needed. The tasks and scheduled time of the recording sessions were decided on in collaboration with the participants, such that the recordings were representative of their normal ADLs while minimizing interruptions to their schedules and preventing recordings of situations that they did not want recorded (e.g. activities with privacy concerns). The recorded tasks included but were not limited to eating or preparing food, performing household tasks (laundry, putting away groceries, using a computer, playing a video game, etc.), or non-private personal tasks (e.g. combing hair, exercise, driving, etc.). Although efforts were made to minimize the presence of other people in the videos, any individual who did appear in segments of the videos (e.g. caregivers) provided informed consent for the study. Figure 1 shows an example of some of the activities in the dataset.

**Figure 1:**
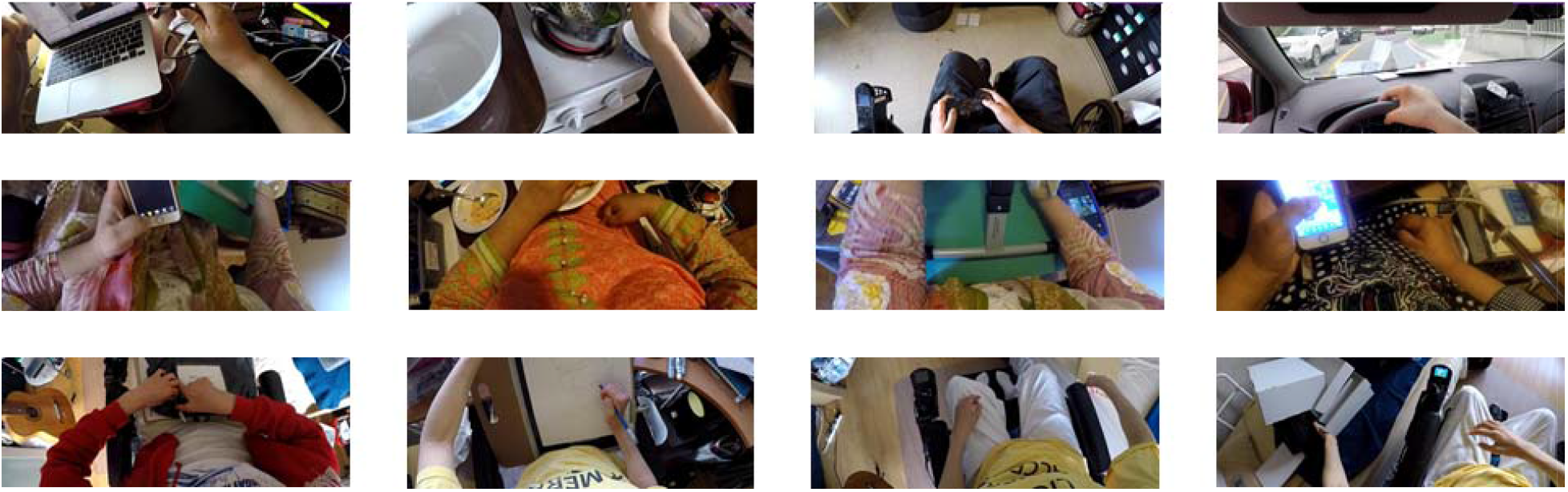
Example frames of the dataset collected for each of the three participants

### Feasibility measures

To characterize the video recordings obtained, we first extracted the total recording time and usable recording time for each participant. The footage was considered usable when the camera was positioned correctly to capture the hands, and when it did not accidentally contain any unwanted recordings (e.g. individuals that did not sign the consent form or footage containing confidential information). Additionally, information relating to participants’ recording behavior was tallied, including number of recording days, number of recording sessions per day, and average duration of recording sessions. Finally, we also compared the number of scheduled activities with the number of activities actually performed in the video recordings.

Beyond tracking the compliance with the planned schedule, we also kept track of any issues that were raised, including troubleshooting with equipment or additional concerns resulting in home visits, scheduling issues that resulted in delayed or missed recordings, as well as any other possible concerns that were reported by the participant.

Structured and semi-structured interviews were administered at the end of the recording session similar to those described in [29]. The questionnaires explored three main topics relating to the use of wearable cameras: privacy, perceived usefulness of the information obtained, and usability. Each section consisted of structured questions on a 5-point Likert scale, as well as an opportunity for open-ended comments. The privacy questions focused on concerns over sharing information from the wearable cameras with different stakeholder groups (i.e. clinicians and researchers), as well as the distinction between sharing raw videos and sharing only metrics automatically extracted from these videos (i.e. measures such as duration and frequency of hand use that would be extracted by a video processing algorithm without the videos being watched by a human). The second set of questions focused on how useful the participants felt the information would be for different stakeholders (i.e. clinicians, researchers, and participants themselves). Lastly, the usability questions focused on factors that might encourage or dissuade participants from using the system. These three categories (privacy, usefulness and usability) were chosen because wearable technology must overcome these challenges to be translated into practice. In addition to the participant interviews, we also interviewed one caregiver, who was a family member of a participant.

The study participants provided written consent prior to participation in the study, which was approved by the Research Ethics Board of the institution (Research Ethics Board, University Health Network: 17-5322).

### Hand use measures

In our previous work in [15] and [16], we demonstrated the possibilities of egocentric video for extracting the interactions of the hand with objects. The system consisted of a binary classification for detecting interaction or no interaction in each video frame, summarized over time.

In the present study, the framework used for hand-object interaction detection consisted of three processing steps, similar to [16]. First, the hand location was determined in the form of a bounding box. This was accomplished using a deep-learning-based object detector (YOLOv2 [30]), re-trained to detect the left and right hands of the user, and the hands of other people in the frame [31]. This method is in contrast to our previous work where hand detection was based on a Regional Convolution Neural Network (R-CNN) [32]. Next, the bounding box was processed for hand segmentation, where the pixels of the hand were separated from the non-hand pixels. With the hand being located and segmented, image features including hand motion, hand shape and colour distribution were extracted for the classification of hand-object interaction. The hand segmentation and interaction detection steps were trained in our previous study, using data from individuals with SCI performing ADLs in the home simulation lab [16].

A total of 108,050 frames from the three participants in the home recordings were labelled and used in the evaluation. The binary classifier trained in [16] was evaluated using data manually labelled by two human observers, where each frame was either classified as interaction or no interaction. Labelling was performed on a frame-by-frame basis, with no bounding box or segmentation shown to the annotator. An annotator was instructed to label the user’s left hand, the user’s right hand, and other people’s hands separately.

## Results

### Feasibility analysis

The feasibility results are summarized into Figure 2. Figure 2a shows the total recording time and usable recording time for each participant. Figure 2b shows the total number of days of the study (from participants receiving the camera to returning the camera) and the number of days during that time in which they collected video. Finally, Figure 2c shows the number of activities performed (i.e. number of distinct activities observed in the video) vs. the number of activities scheduled (i.e. the number of distinct activities planned with the participants during the first appointment).

All three participants performed at most one recording session in a given day with an average recording duration of 32±5 minutes per session. Two out of the three participants completed the study later than the planned 21 days, due to technical issues with the devices and conflicts in personal schedules (e.g. health, family matter, etc.). Technical issues relating to operation of the camera resulted in an additional home visit for two participants.

**Figure 2:**
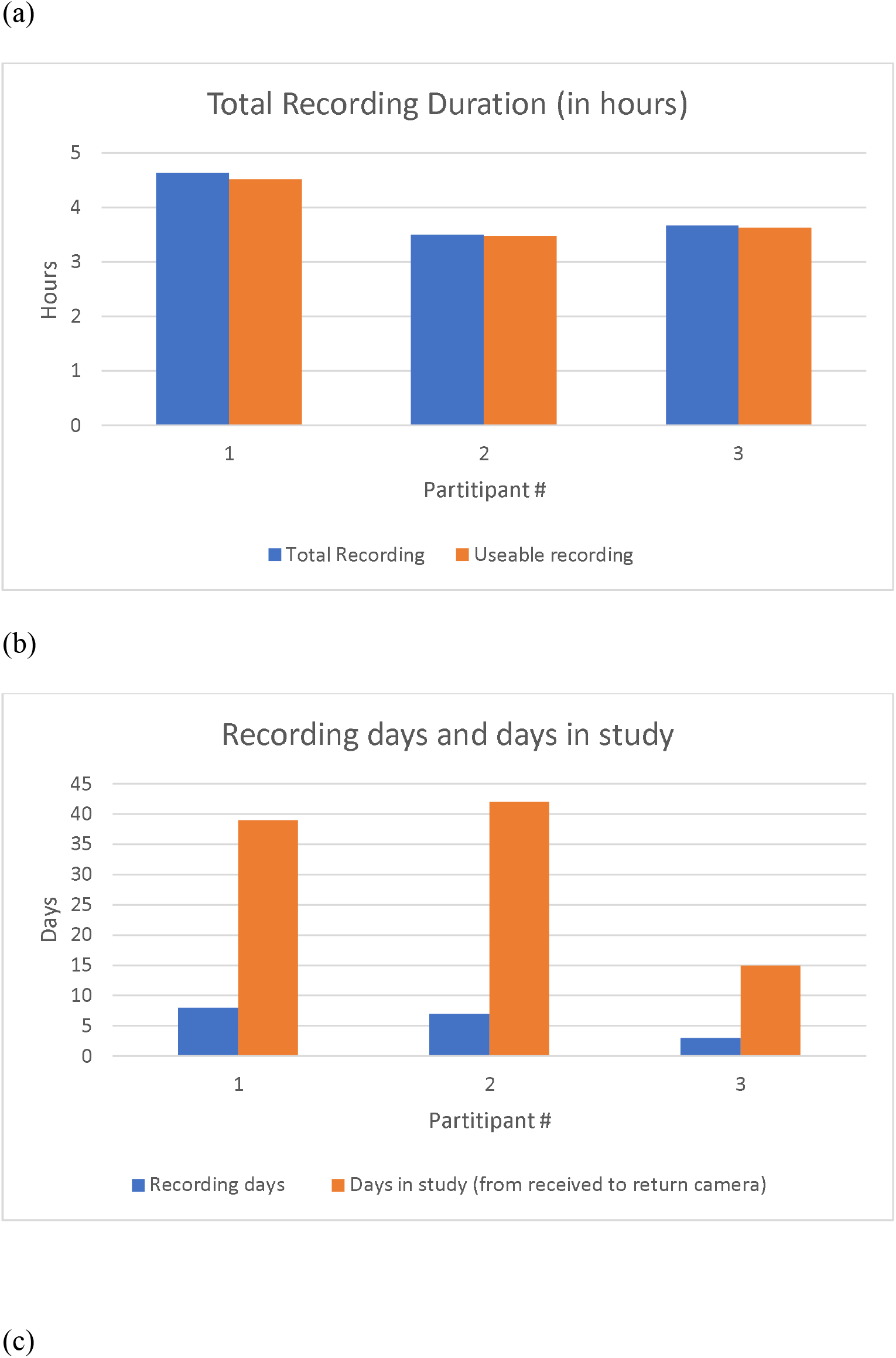
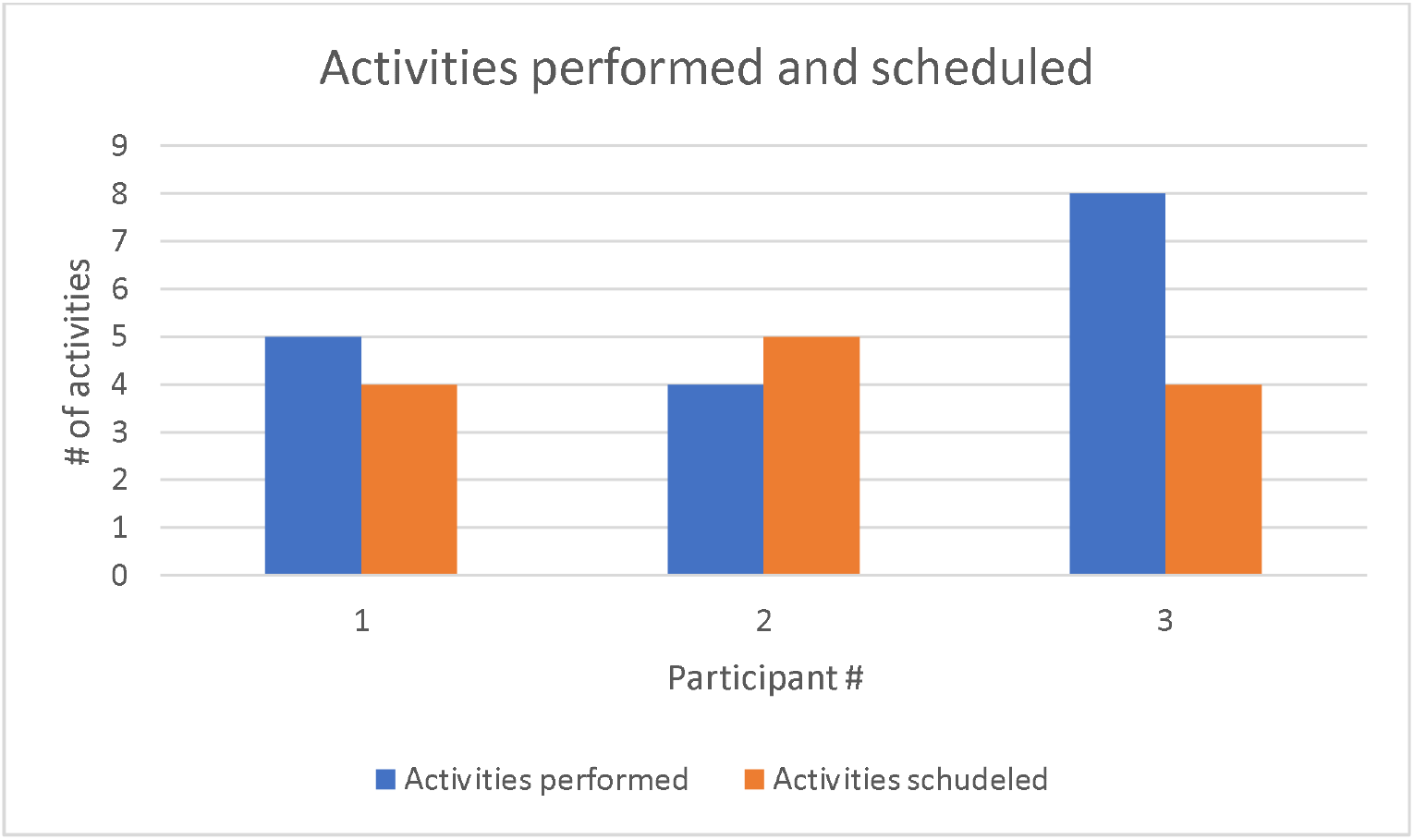
Feasibility metrics regarding video data obtained. (a) Total recording time and total usable recording time for each participant, (b) number of recording days and the number of days in the study, from participant receiving the equipment to returning the equipment, and (c) number of activities obtained compared to the number of activities planned.

### Participant feedback

The questions regarding privacy for each participant of this study are shown in Figure 3a. Participants in this study expressed relatively low levels of concern about egocentric video being stored and used by clinicians or by researchers, with a mean of 1.33±0.58 in both cases. When asked about storing only automatically extracted metrics rather than the videos themselves, all participants reduced these concerns down to 1.00±0.00 (the lowest possible score) regardless of whether the data was to be used by clinicians or researchers. Nevertheless, privacy concerns were slightly raised when asked to wear a camera in daily life at home (1.33±0.58). The reason provided by the individual was the potential risk of ascertaining their location or leaking private information, as well as the inconvenience of concealing unwanted footage, e.g. having to cover the password on the computer screen. They described little concerns about other family members appearing in the video (1.33±0.15). Participants were somewhat comfortable with wearing the camera at home (3.33±1.55) and in public (3.67±1.55). However, they noted that capturing a public recording of other people without permission could cause conflict and unwanted attention. When it comes to the control of the devices, all participants saw a relatively high value in the ability to start and stop recording at any time (4.33±1.15) as well as ability to review the video (4.00±1.00).

**Figure 3:**
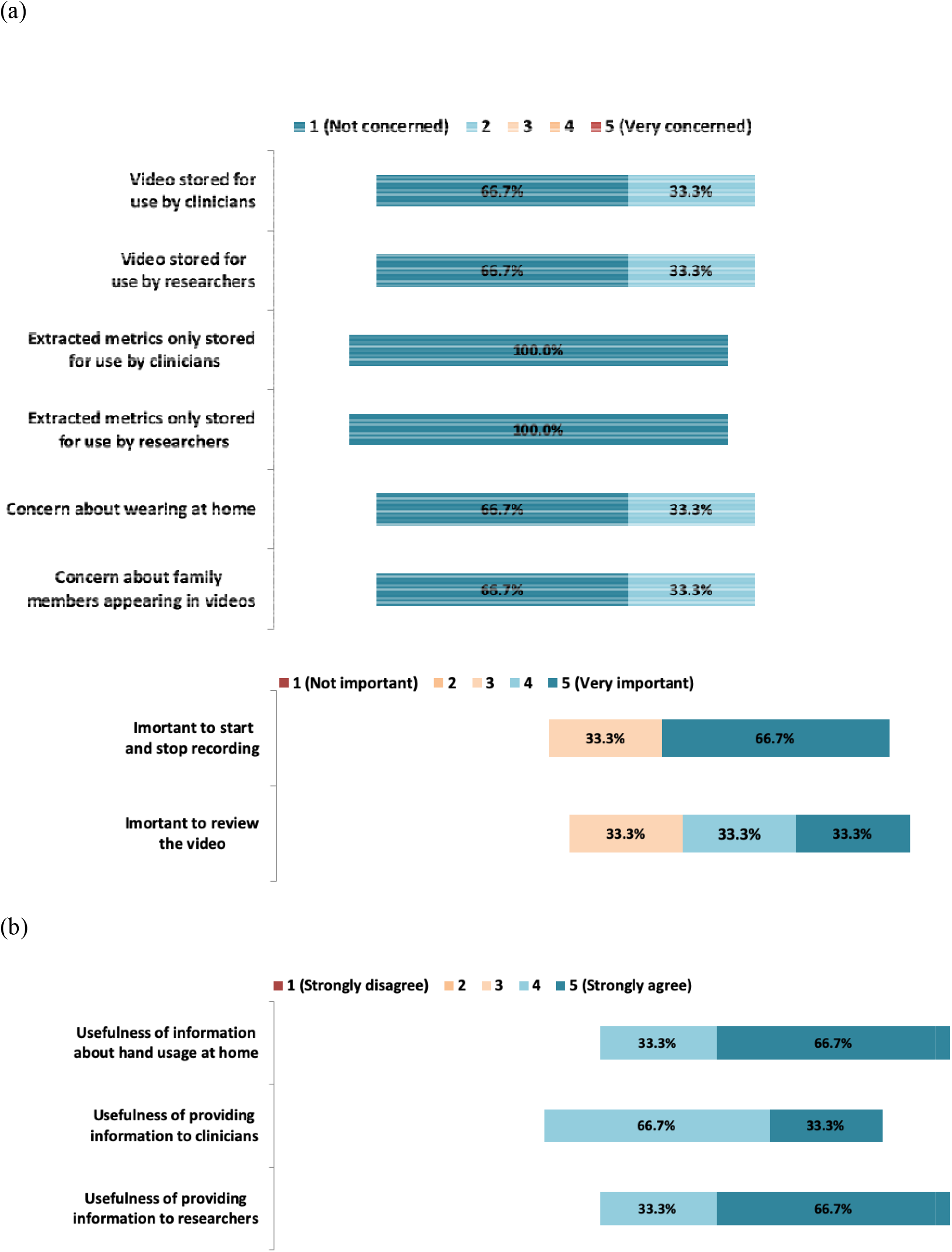
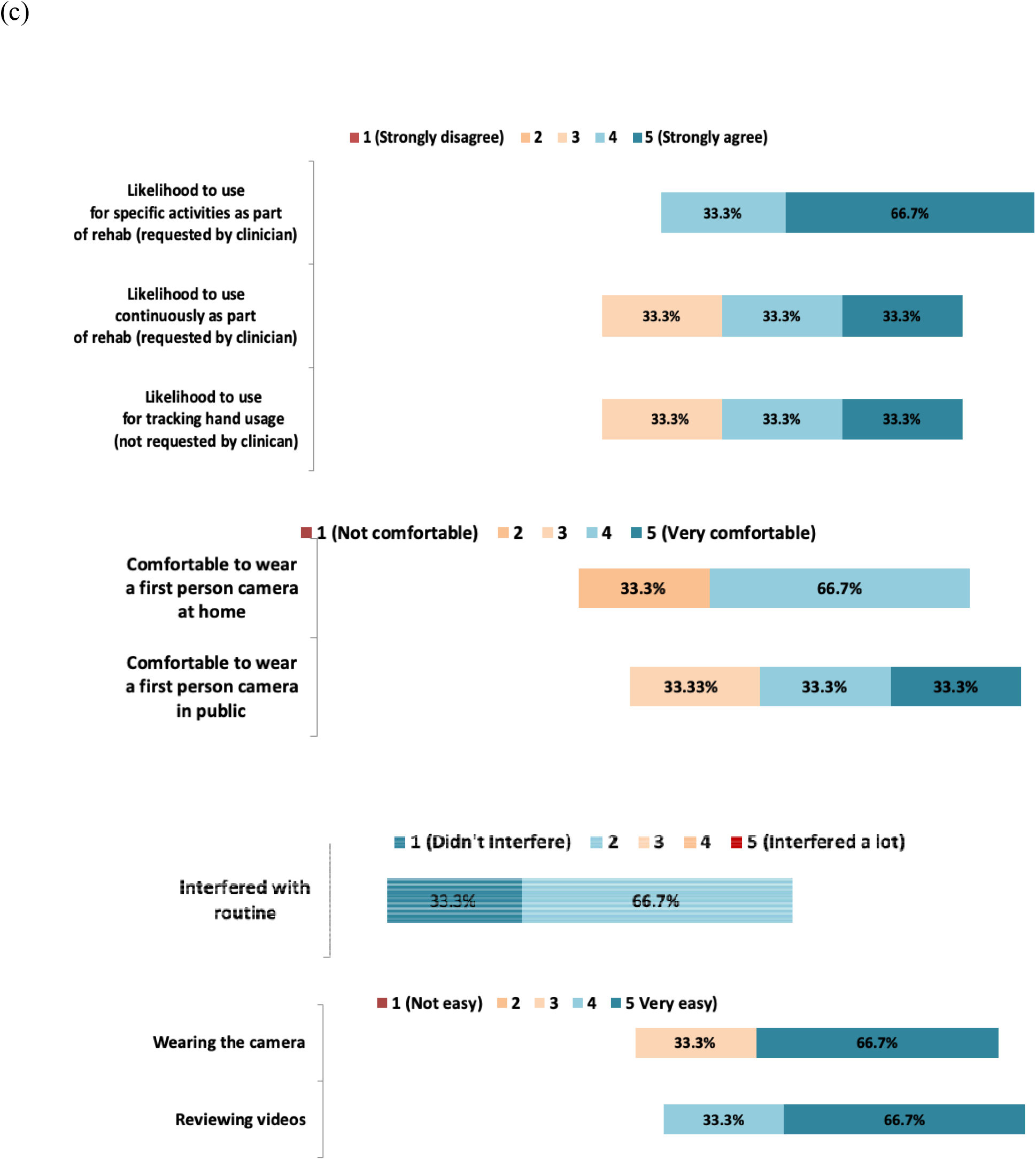
Distribution of answers to questions pertaining to: (a) privacy of information, (b) usefulness of the information, and (c) usability of the technology.

In regard to the usefulness of the information collected (Figure 3b), participants believed that the hand usage information collected from an egocentric camera in their home would be useful to clinicians (4.33±0.58) and more so for researchers (4.67±0.58). Participants also saw the value in keeping track of their own hand function recovery (4.67±0.58).

For usability (Figure 3c), we explored how often participants would comply with the use of the camera. They reported high likelihood of using an egocentric device to record specific activities during their rehabilitation process, when prescribed by a doctor or therapist (4.67±0.58). However, this level of compliance dropped in a case where a doctor or therapist asked them to wear the camera all the time during their rehabilitation process (4.00±1.00). This drop-in compliance continued at the same level when the participants were asked if they were willing to use the wearable camera to track their hand usage for their own use (i.e. without medical instruction or involvement). One participant stated that the current GoPro camera can feel a little heavy over time. Nevertheless, they still felt that the camera is somewhat easy to use and not difficult to set up (4.33±1.15) and that it was easy to review the video before sharing with the researchers (4.33±0.57). However, according to one of the participants, this review process can take some time because of the long video footage. Nevertheless, the overall consensus was that the video recording did very little to interfere with their daily routine (1.33±0.58).

When we asked participants how many hours per week they would be willing to wear the system as part of their rehabilitation, they indicated that they were willing to record more often if asked to do so only for a short term (participants denoted short term as about one month of 3 hours per week, no more than 7 hours a week, with 1-2 hours per session. A shorter duration of recording per week was preferred (i.e. <1 hours). For longer-term recording (i.e. >1 month), the participants indicated that recording sessions and hours per week should be shorter (e.g.1 hour per week with sessions ranging from 10 - 30 minutes.)

The caregiver expressed a low level of concern (1.0, lowest possible score) about appearing in video that is intended for assessing hand function by a clinician or a researcher. Again, an identical score was reported when the caregiver was asked if they had any concerns about a wearable camera system that does not store recorded video but uses a computer algorithm to analyze the video and output hand function data. Additional questions relating to a household member wearing a camera in daily life at home and in public also revealed no concerns by the caregiver. In the open-ended section, the caregiver indicated that he/she did help the household member in using the camera system. The caregiver assisted the participant in putting on the camera head strap, replacing the battery or accessing SD card for the purposes of viewing the video on the computer. The assistance required was often short and lasted around 30 seconds.

### Functional Hand use

Finally, we evaluated the performance of the hand-object interaction detector using the collected home video data. Table 2 shows the F1-score and accuracy where the predicted interactions from the algorithm output are compared with the actual interactions from the manually labelled data. The processing time of the entire hand-object interaction detection pipeline is 0.21 second per frames from the input image frame to the output metrics using an Intel-i7-8700k, DDR4-16GB, GTX1080Ti-11GB, and Ubuntu16.04 LTS (64-bit). The mean F1-score and accuracy for all three participants (left and right hand) are 0.75 ± 0.08 and 0.72 ± 0.19, respectively.

**Table 2:** F1-score and accuracy of the hand-object interaction for left (L) and right (R) hand for each of the participants.

|  | F1-score |  | Accuracy |  |
| --- | --- | --- | --- | --- |
| Participants | L | R | L | R |
| 1 | 0.59 | 0.87 | 0.74 | 0.88 |
| 2 | 0.84 | 0.56 | 0.76 | 0.47 |
| 3 | 0.82 | 0.82 | 0.75 | 0.73 |
| Mean ± S.D. | $0.75 \pm 0.14$ | $0.75 \pm 0.16$ | $0.75 \pm 0.01$ | $0.69 \pm 0.21$ |

## Discussion

This study evaluated the feasibility of deploying an egocentric wearable camera to capture the hand use of individuals with SCI in their home environment. The information from this study is needed because limited methods are available for capturing rehabilitation progress in the community, and accurate outcome measures are critical for the advancement of new interventions. We deployed the wearable cameras in the community, where participants were instructed to record their ADLs at home without the presence of researchers. Using this data, we showed that it is possible to capture egocentric recordings of ADLs at home. The study provides feasibility information necessary to the planning of egocentric video recording in the community. Furthermore, hand use in ADLs was automatically quantified from the egocentric videos using a custom algorithmic framework. The proposed wearable system was deployed in the real world for the first time. The information obtained may help to form the basis for new home-based outcome measures that reflect performance of UE tasks, and thus give insight into independence.

In evaluating feasibility, we found that the majority of recordings had a view of the hand and were thus usable for capturing hand-use information. Unfortunately, two out of three participants took more than the planned 21 days to complete the study. This delay was attributable to technical issues with the devices and conflict in personal schedules. While most participants managed to meet or even exceed the activities planned with the researcher, the activities were not diverse and most of the recordings have repetition across recorded sessions and days. The lack of diversity is concerning because other ADLs might be missed, and so a clearer scheduling plan is required. In another words, the recording schedule needs to consider a diversity of activities while still maintaining the flexibility of recording time for the participants.

Since the participants expressed a preference for short recordings (i.e. 12 - 39 minutes), we recommend emphasizing recordings lasting approximately 1 hour. This will enable meaningful capture of hand use in a natural context while keeping the recording relatively short. Participants may need to keep the recording devices for at least 30 days rather than for just 21 days. These extra days would leave room for unforeseen issues to be addressed and for participants to record more unique activities. Clear communication with the participants early on is necessary to ensure that the videos genuinely reflect their normal routine, rather than artificial activities with more hand use than usual.

The performance of the hand-object interaction detection was comparable here to the results reported in a home simulation laboratory (e.g. F1-scores of 0.75 ± 0.14 vs. 0.74 ± 0.15 for the left hand and 0.75 ± 0.16 vs. 0.73 ± 0.15 for the right hand, respectively) [16]. This consistency demonstrates the potential of a wearable egocentric camera for capturing quantitative measures of hand use as well as the robustness and generalizability of the system in the home environment.

In this study, we established the feasibility of deploying a wearable camera system to track functional hand use after SCI in the target setting, the home. We additionally found that the technical performance obtained in the laboratory was transferable to the less constrained real home environment. With proper recording scheduling, we believe that such a tool will provide clinicians and researchers with a basis for objective measures of hand function in the home. This framework lays the groundwork for studies to further validate new egocentric video-based UE outcomes after SCI.

## Data Availability

N/A

## Acknowledgments

This study was supported in part by the Natural Sciences and Engineering Research Council of Canada (RGPIN-2014-05498), the Rick Hansen Institute (G2015-30), and the Ontario Early Researcher Award (ER16-12-013).

The authors would like to thank Gregory Wong for his valuable assistance in the data labelling process. The authors would also like to thank all the participants of the study.

## Notes

### Competing Interest Statement

The authors have declared no competing interest.

### Author Declarations

The study participants provided written consent prior to participation in the study, which was approved by the Research Ethics Board of the institution (Research Ethics Board, University Health Network, Canada: 17-5322).

